# Virtual Therapy Habit Reversal Training for Body-Focused Repetitive Behaviors: Clinical Outcomes from a Large Real-World Sample of Youth and Adults

**DOI:** 10.1101/2025.03.10.25323675

**Authors:** Jamie D. Feusner, Clare C. Beatty, Christopher Murphy, Patrick B. McGrath, Nicholas R. Farrell, Mia Nuñez, Nicholas Lume, Reza Mohideen, Larry Trusky, Stephen M. Smith, Andreas Rhode

## Abstract

**Objective:** To examine the effectiveness of virtual therapy-delivered habit reversal training (HRT) in large real-world samples of children, adolescents, and adults with trichotillomania and excoriation disorder.

**Methods:** The sample included 543 patients with trichotillomania (57 children, 75 adolescents, 408 adults) and 528 patients with excoriation disorder (40 children, 46 adolescents, 442 adults). Treatment followed a protocol of twice-weekly HRT sessions, transitioning to weekly sessions. The Repetitive Body Focused Behavior Scale (RBFBS) was administered at baseline, weeks 5-7, weeks 14-16, and during maintenance periods through week 52.

**Results:** At weeks 14-16, trichotillomania showed a median 33.33% severity reduction (IQR=11.11%-54.55%; 44.08% achieving ≥35% reduction) with large effects (Hedges’ g=1.01, 95% CI [0.88, 1.14]). Excoriation showed a median 33.33% reduction (IQR=12.50%-57.14%; 48.66% achieving 35% reduction; g=1.16 [1.02-1.30]). Improvements were maintained through week 52 (trichotillomania: g=1.51 [CI: 1.23 to 1.79]; excoriation: g=1.56 [1.29-1.84]). Both conditions showed improvements in depression, anxiety, and stress (g=0.22-0.29). Mean treatment duration was 14.64±2.50 weeks (7.71±2.61 sessions) for trichotillomania and 14.54±2.69 weeks (7.73±2.68 sessions) for excoriation. All age groups improved, with effect sizes ranging from g=0.78-1.11 for trichotillomania and g=0.68-1.54 for excoriation.

**Conclusion:** This analysis demonstrates that virtual therapy-delivered HRT can effectively reduce both hair-pulling and skin-picking severity and improve related symptoms in a real-world setting. The large treatment effects and improvements across the lifespan for both conditions suggest this delivery format may help address barriers to accessing evidence-based care for body-focused repetitive behaviors.

---

Body-focused repetitive behaviors (BFRBs) are psychiatric conditions characterized by repetitive, ritualized behaviors directed at one’s body, involving compulsively damaging one’s physical appearance or causing physical injury ^1^. While occasional engagement in these behaviors appears common in the general population, pathological forms causing significant distress and functional impairment affect approximately 12% of individuals ^2^. These conditions are associated with elevated rates of anxiety and diminished quality of life ^3^. The two most extensively studied BFRBs, now recognized as distinct disorders within the DSM-5’s Obsessive-Compulsive and Related Disorders category, are trichotillomania and excoriation disorder^4^.

Trichotillomania is characterized by recurrent hair pulling, leading to noticeable hair loss and functional impairment ^4^. The first large-scale U.S. epidemiological survey found a point prevalence of 1.7% among adults ^5,6^, aligning with lifetime prevalence estimates of ∼1-3% ^7–12^. Trichotillomania typically onsets during early adolescence, between ages 10-12 ^5,6,13,14^ and frequently co-occurs with anxiety and depression ^10,15^. Despite its impact on quality of life, approximately 65% of affected individuals never seek treatment ^16^.

Excoriation disorder (ED) is characterized by recurrent skin picking, leading to tissue damage and functional impairment (American Psychiatric Association, 2013). Prevalence estimates range from 1.4% to 5.4% in the general population ^17,18^, with a recent large-scale study finding that 2.1% of adults meet diagnostic criteria for the condition ^6,19^. Excoriation disorder typically onsets during adolescence ^20^ and frequently co-occurs with depression, anxiety, and obsessive-compulsive disorder, which may contribute to increased symptom severity ^21,22^. The condition often leads to significant physical scarring ^19^ and psychosocial impairment, including missed work and social obligations ^23,24^, yet like trichotillomania, many affected individuals do not seek mental health treatment.

For those who do seek treatment, Habit Reversal Training (HRT), a form of cognitive-behavioral therapy, is an effective form of treatment for BFRBs ^25–27^. HRT helps patients develop awareness of picking and pulling behaviors and replace them with competing responses ^28–30^, incorporating self-monitoring, awareness training, competing response training, and stimulus control procedures.

HRT controlled studies have shown large effects (ES =-1.14), and superiority to pharmacological interventions ^26^ and controls ^31,32^. Research also supports the effectiveness of HRT when enhanced with components from acceptance and commitment therapy (ACT)^33^ and dialectical behavior therapy (DBT) components^34^. Evidence supports HRT’s efficacy in youth with trichotillomania, with studies showing significant symptom reduction compared to control conditions^35^ and superior outcomes (76% vs 21%, d =1.31; response rate) compared to treatment as usual, where youth continued their existing treatments including psychotherapy and medications ^36^. Evidence also supports HRT’s efficacy for excoriation disorder, with randomized controlled trials demonstrating symptom reduction ^37,38^, though primarily in short-term outcomes. While HRT demonstrates efficacy in controlled trials and generally maintains gains for 3-6 months post-treatment, many individuals do not respond or experience relapse, with 50-67% of initial treatment responders relapsing during long-term follow-up ^39^.

Despite the established efficacy of HRT, multiple barriers impede access to treatment for both trichotillomania and excoriation disorder. Provider limitations represent a significant challenge - a survey of healthcare professionals found that while they possessed basic trichotillomania knowledge, many endorsed non-empirically supported treatments and lacked resources for patients ^40,41^. This knowledge gap is reflected in practice, with only 3% of hair pullers perceiving their providers as trichotillomania experts ^16^ and more than a quarter rating their providers as “not at all knowledgeable” about the condition. Recent studies continue to demonstrate that providers working with trichotillomania patients often lack sufficient knowledge about both the condition and its treatment options ^40^ Similar challenges exist for excoriation disorder, where many affected individuals seek dermatologic rather than mental health care due to limited provider expertise ^42^. Geographic and cost barriers further compound these challenges, particularly affecting individuals in areas without specialized mental health services.

The rise of telehealth platforms offers a promising solution to expand access to BFRB treatment while addressing many traditional barriers. Virtual care delivery has become increasingly integrated into mental health services, potentially allowing patients to connect with knowledgeable providers regardless of geographic location. Emerging research on technology-based interventions for BFRBs has shown encouraging results, with studies demonstrating effectiveness of virtually-delivered treatments in both provider-led interventions ^43,44^ and automated approaches ^45^ for trichotillomania. Initial studies examining telehealth delivery of acceptance-enhanced behavior therapy (AEBT), which integrates traditional HRT with acceptance and commitment therapy (ACT), have shown significant reductions in hair-pulling severity, with large effects observed for both adults (ω²=.473)^43^ and adolescents (Hedges’ g=1.55)^44^, with high patient satisfaction reported. However, research suggests that telehealth delivery of HRT may result in differential outcomes compared to in-person treatment, with factors such as technology familiarity and limited field of view potentially impacting effectiveness ^46^. This consideration may be particularly relevant for excoriation disorder, where visual assessment of skin-picking behaviors and resulting damage is crucial for treatment monitoring ^46^.

While these initial clinical trials of remote HRT delivery are promising, research has not yet examined outcomes in large-scale, real-world clinical settings. NOCD, a virtual behavioral health provider specializing in obsessive-compulsive and related disorders, delivers HRT through virtual therapy with HRT-trained therapists, supplemented by between-session messaging and an online community platform. This study aimed to determine clinical outcomes in a large naturalistic sample of individuals with trichotillomania or excoriation disorder who received HRT via NOCD’s online specialty therapy platform, addressing a critical gap in understanding the real-world effectiveness of technology-enabled remote treatment delivery.

## Method

### Sample

This was a retrospective, observational longitudinal analysis of clinical data from individuals who received HRT treatment for trichotillomania or excoriation disorder through NOCD’s online specialty therapy platform between May 2021 and February 2025. We applied several inclusion criteria to the data that were included for both conditions: (1) valid baseline RBFBS assessment, (2) baseline RBFBS score > 0, (3) completion of ≥2 assessments, (4) completion of at least one follow-up assessment (weeks 5-7 OR weeks 14-16), and (5) a minimum of five treatment sessions. The final analysis included 543 trichotillomania patients (51.2% of 1,060 initial patients) and 528 excoriation disorder patients (43.2% of 1,222 initial patients)), including children, adolescents, and adults with primary diagnoses of their respective conditions.

### Initial Evaluation and Clinical Assessments

Initial evaluations were conducted by NOCD-trained therapists using the Diagnostic Interview for Anxiety, Mood, and Obsessive-Compulsive and Related Neuropsychiatric Disorders (DIAMOND; ^4,47^). Patients who met DSM-5 criteria for trichotillomania or excoriation disorder were eligible for treatment. Those rated as having “extreme” symptom severity were typically referred to more intensive treatment options (see Supplementary Material).

### Treatment Approach

The NOCD treatment model delivered HRT through a structured virtual therapy. The initial phase included twice-weekly 60-minute video sessions for 2-3 weeks for most, followed by 10-14 weeks of weekly 60-minute sessions. Some transitioned to 30-minute check-in sessions based on clinical progress, symptom reduction, and mastery of HRT principles. All sessions used HIPAA-compliant video conferencing. Patients also had access to in-app messaging with their therapist, a mobile app for tracking, and a moderated online community. Treatment involved core HRT components (awareness training, competing response training, generalization training, and social supports^48^) and occasionally incorporated stimulus control strategies and functional analysis of the BFRBs^49,50^. Treatment was delivered by therapists who completed intensive training in OCD-related disorders and HRT (see Supplementary Material for detailed training procedures). The treatment duration and session frequency were adjusted based on individual needs.

### Assessments

While patients completed assessments every three weeks throughout treatment, we analyzed outcomes at three primary intervals: baseline, weeks 5-7 (to capture early treatment response), and weeks 14-16 (representing the primary endpoint). Additional assessments collected during follow-up periods (weeks 17-52) were analyzed to evaluate long-term outcomes (see Supplementary Material for detailed assessment timeline and procedures).

The primary outcome measure was the Repetitive Body Focused Behavior Scale (RBFBS^51^) administered in both self-report (RBFBS-SR) and parent-report (RBFBS-P) formats. The RBFBS assesses three types of body-focused repetitive behaviors (skin picking, hair pulling, and nail-biting) using 12 items. Respondents rate three domains on a 0-4 scale: time spent, interference, and distress (emotional impact, urge resistance). Hair-pulling and skin-picking subscale scores range from 0-12 (mild: 1-3, moderate: 4-6, severe: 7-9, extreme: 10-12). The parent-report version shows excellent internal consistency for hair pulling (α=.95) and skin picking (α=.93) subscales ^51^, while adult data supports convergent validity with established measures (r=.74 with GBS-8)^52–54^.

Secondary measures assessed depression, anxiety, and stress (Depression, Anxiety, and Stress Scale; DASS-21^55^), quality of life (Quality of Life Enjoyment and Satisfaction Questionnaire-Short Form; Q-LES-Q-SF^56^), and disability and functioning (World Health Organization Disability Assessment Schedule 2.0; WHODAS 2.0^57^). See Supplementary Material for complete measure descriptions.

### Data Analysis

Treatment engagement metrics (messaging, app opens, visits) were calculated through week 16, analyzing message frequency and app usage patterns. Primary and secondary outcomes were analyzed using linear mixed models with assessment timepoint as a fixed factor, patient as a random factor, and respective outcome scores (BFRB severity, depression, anxiety, stress, quality of life, and disability) as dependent variables. Age effects were examined by testing interactions between age group and evaluation timepoints.

Treatment response categories (≥45% improvement, ≥35% improvement, ≥25%, 0-24%, or worsened ^58^) were based on meta-analyses of trichotillomania clinical trials ^59–61^. Effect sizes were calculated using Hedges’ g with 95% confidence intervals. Treatment duration effects were examined by comparing outcomes between shorter (14-18 weeks) and longer (30-36 weeks) durations. Analyses were conducted using R Statistical Software (v4.1.3). Detailed information about missing data and satisfaction analyses is provided in Supplementary Material.

### Ethical Considerations

The analyses in this study did not require research ethics board review as this does not meet the criteria for Human Subject Research as defined by federal regulations for human subject protections, 45 CFR 46.102(e); this is a secondary analysis of de-identified data from clinical records, obtained and analyzed retrospectively, and was not the result of a research intervention or interaction. Compliance with data protection laws was ensured through NOCD’s privacy policy, which all patients agreed to, outlining data use and protection measures.

## Results

### Sample

The analysis included 1,071 eligible patients with either trichotillomania or excoriation disorder (see Table 1 for detailed demographics). Both conditions showed a predominance of adult patients, though the age distribution differed between groups. The trichotillomania sample was 75.6% adult (mean age =31.0 years, SD =8.96), with 13.9% adolescents (mean age =14.9 years, SD =1.41) and 10.6% children (mean age =10.2 years, SD =1.83). The excoriation disorder sample was 83.7% adult (mean age =30.7 years, SD =9.87), with 8.7% adolescents (mean age =15.6 years, SD =1.22) and 7.6% children (mean age =10.5 years, SD =1.62).

**Table 1.**
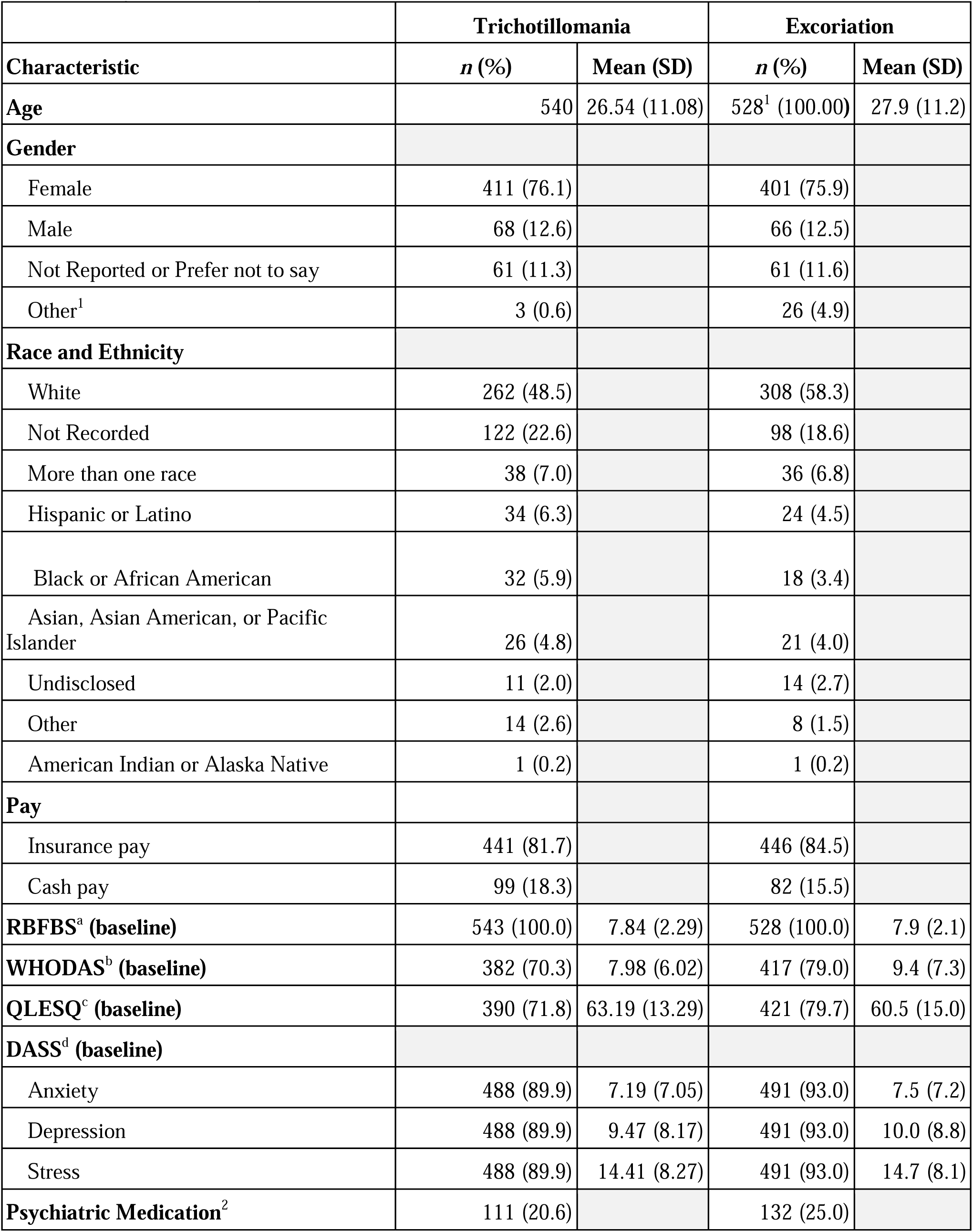
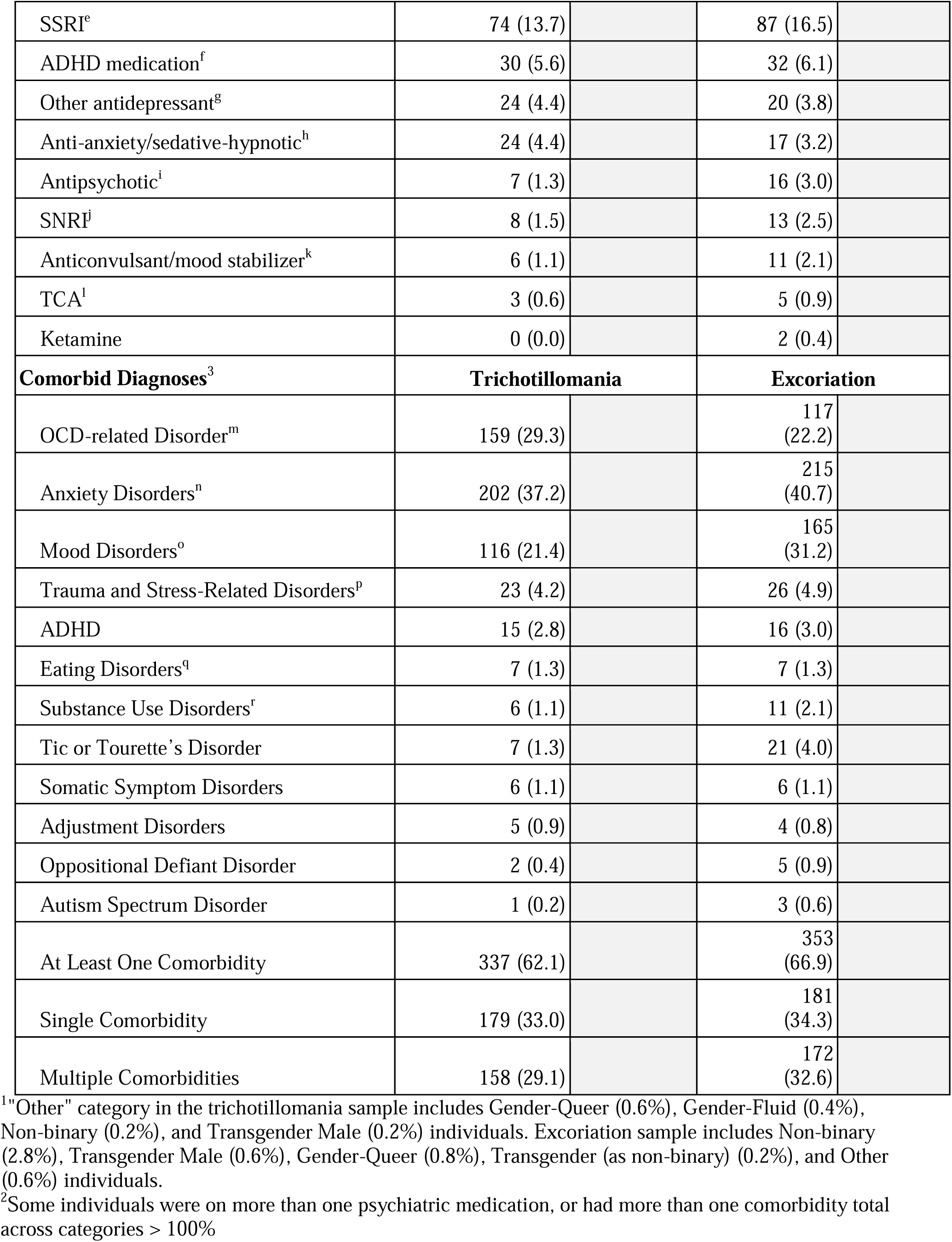
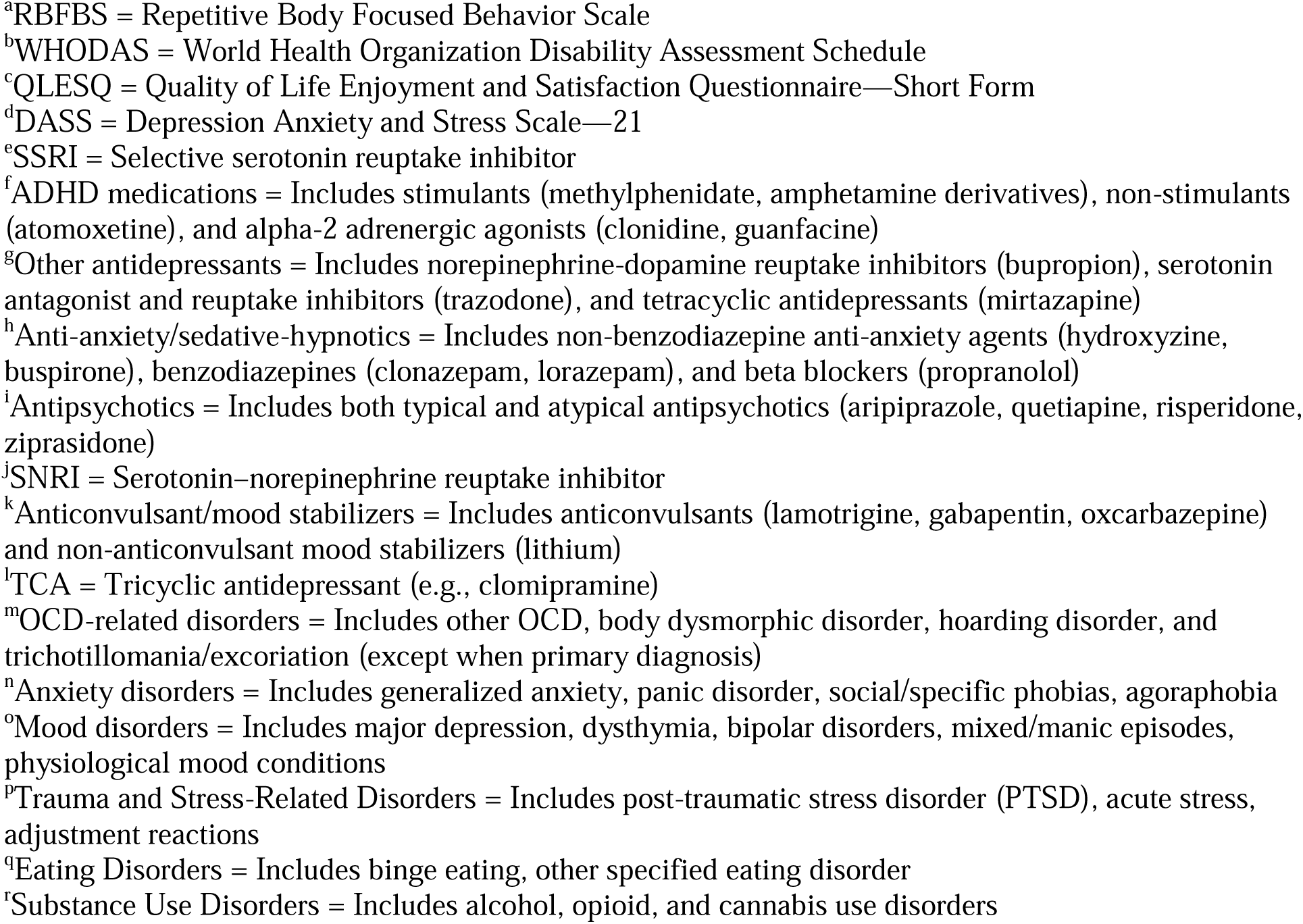
Demographics and psychometrics.

|  | **Trichotillomania** |  | **Excoriation** |  |
| --- | --- | --- | --- | --- |
| **Characteristic** | ***n* (%)** | **Mean (SD)** | ***n* (%)** | **Mean (SD)** |
| **Age** | 540 | 26.54 (11.08) | 528<sup>1</sup> (100.00) | 27.9 (11.2) |
| **Gender** |  |  |  |  |
| Female | 411 (76.1) |  | 401 (75.9) |  |
| Male | 68 (12.6) |  | 66 (12.5) |  |
| Not Reported or Prefer not to say | 61 (11.3) |  | 61 (11.6) |  |
| Other<sup>1</sup> | 3 (0.6) |  | 26 (4.9) |  |
| **Race and Ethnicity** |  |  |  |  |
| White | 262 (48.5) |  | 308 (58.3) |  |
| Not Recorded | 122 (22.6) |  | 98 (18.6) |  |
| More than one race | 38 (7.0) |  | 36 (6.8) |  |
| Hispanic or Latino | 34 (6.3) |  | 24 (4.5) |  |
| Black or African American | 32 (5.9) |  | 18 (3.4) |  |
| Asian, Asian American, or Pacific Islander | 26 (4.8) |  | 21 (4.0) |  |
| Undisclosed | 11 (2.0) |  | 14 (2.7) |  |
| Other | 14 (2.6) |  | 8 (1.5) |  |
| American Indian or Alaska Native | 1 (0.2) |  | 1 (0.2) |  |
| **Pay** |  |  |  |  |
| Insurance pay | 441 (81.7) |  | 446 (84.5) |  |
| Cash pay | 99 (18.3) |  | 82 (15.5) |  |
| **RBFB<sup>a</sup> (baseline)** | 543 (100.0) | 7.84 (2.29) | 528 (100.0) | 7.9 (2.1) |
| **WHODAS<sup>b</sup> (baseline)** | 382 (70.3) | 7.98 (6.02) | 417 (79.0) | 9.4 (7.3) |
| **QLESQ<sup>c</sup> (baseline)** | 390 (71.8) | 63.19 (13.29) | 421 (79.7) | 60.5 (15.0) |
| **DASS<sup>d</sup> (baseline)** |  |  |  |  |
| Anxiety | 488 (89.9) | 7.19 (7.05) | 491 (93.0) | 7.5 (7.2) |
| Depression | 488 (89.9) | 9.47 (8.17) | 491 (93.0) | 10.0 (8.8) |
| Stress | 488 (89.9) | 14.41 (8.27) | 491 (93.0) | 14.7 (8.1) |
| **Psychiatric Medication<sup>2</sup>** | 111 (20.6) |  | 132 (25.0) |  |
| SSRI<sup>e</sup> | 74 (13.7) |  | 87 (16.5) |  |
| ADHD medication<sup>f</sup> | 30 (5.6) |  | 32 (6.1) |  |
| Other antidepressant<sup>g</sup> | 24 (4.4) |  | 20 (3.8) |  |
| Anti-anxiety/sedative-hypnotic<sup>h</sup> | 24 (4.4) |  | 17 (3.2) |  |
| Antipsychotic<sup>i</sup> | 7 (1.3) |  | 16 (3.0) |  |
| SNRI<sup>j</sup> | 8 (1.5) |  | 13 (2.5) |  |
| Anticonvulsant/mood stabilizer<sup>k</sup> | 6 (1.1) |  | 11 (2.1) |  |
| TCA<sup>l</sup> | 3 (0.6) |  | 5 (0.9) |  |
| Ketamine | 0 (0.0) |  | 2 (0.4) |  |
| **Comorbid Diagnoses<sup>3</sup>** | **Trichotillomania** |  | **Excoriation** |  |
| OCD-related Disorder<sup>m</sup> | 159 (29.3) |  | 117 (22.2) |  |
| Anxiety Disorders<sup>n</sup> | 202 (37.2) |  | 215 (40.7) |  |
| Mood Disorders<sup>o</sup> | 116 (21.4) |  | 165 (31.2) |  |
| Trauma and Stress-Related Disorders<sup>p</sup> | 23 (4.2) |  | 26 (4.9) |  |
| ADHD | 15 (2.8) |  | 16 (3.0) |  |
| Eating Disorders<sup>q</sup> | 7 (1.3) |  | 7 (1.3) |  |
| Substance Use Disorders<sup>r</sup> | 6 (1.1) |  | 11 (2.1) |  |
| Tic or Tourette's Disorder | 7 (1.3) |  | 21 (4.0) |  |
| Somatic Symptom Disorders | 6 (1.1) |  | 6 (1.1) |  |
| Adjustment Disorders | 5 (0.9) |  | 4 (0.8) |  |
| Oppositional Defiant Disorder | 2 (0.4) |  | 5 (0.9) |  |
| Autism Spectrum Disorder | 1 (0.2) |  | 3 (0.6) |  |
| At Least One Comorbidity | 337 (62.1) |  | 353 (66.9) |  |
| Single Comorbidity | 179 (33.0) |  | 181 (34.3) |  |
| Multiple Comorbidities | 158 (29.1) |  | 172 (32.6) |  |

<sup>1</sup>"Other" category in the trichotillomania sample includes Gender-Queer (0.6%), Gender-Fluid (0.4%), Non-binary (0.2%), and Transgender Male (0.2%) individuals. Excoriation sample includes Non-binary (2.8%), Transgender Male (0.6%), Gender-Queer (0.8%), Transgender (as non-binary) (0.2%), and Other (0.6%) individuals.
<sup>2</sup>Some individuals were on more than one psychiatric medication, or had more than one comorbidity total across categories > 100%
<sup>a</sup>RFBFS = Repetitive Body Focused Behavior Scale
<sup>b</sup>WHODAS = World Health Organization Disability Assessment Schedule
<sup>c</sup>QLESQ = Quality of Life Enjoyment and Satisfaction Questionnaire—Short Form
<sup>d</sup>DASS = Depression Anxiety and Stress Scale—21
<sup>e</sup>SSRI = Selective serotonin reuptake inhibitor
<sup>f</sup>ADHD medications = Includes stimulants (methylphenidate, amphetamine derivatives), non-stimulants (atomoxetine), and alpha-2 adrenergic agonists (clonidine, guanfacine)
<sup>g</sup>Other antidepressants = Includes norepinephrine-dopamine reuptake inhibitors (bupropion), serotonin antagonist and reuptake inhibitors (trazodone), and tetracyclic antidepressants (mirtazapine)
<sup>h</sup>Anti-anxiety/sedative-hypnotics = Includes non-benzodiazepine anti-anxiety agents (hydroxyzine, buspirone), benzodiazepines (clonazepam, lorazepam), and beta blockers (propranolol)
<sup>i</sup>Antipsychotics = Includes both typical and atypical antipsychotics (aripiprazole, quetiapine, risperidone, ziprasidone)
<sup>j</sup>SNRI = Serotonin–norepinephrine reuptake inhibitor
<sup>k</sup>Anticonvulsant/mood stabilizers = Includes anticonvulsants (lamotrigine, gabapentin, oxcarbazepine) and non-anticonvulsant mood stabilizers (lithium)
<sup>l</sup>TCA = Tricyclic antidepressant (e.g., clomipramine)

### Missing Data

Assessment completion patterns were similar between conditions. For trichotillomania, 77.7% provided weeks 14-16 assessments. For excoriation disorder, 77.5% provided weeks 14-16 assessments. Among participants who did not provide week 14-16 assessments, the median treatment duration was 11 weeks [IQR: 9-12] for trichotillomania and 10 [IQR: 9-12] weeks for excoriation disorder. More information can be found in Table 3 and Supplementary Results.

### Overall Treatment Effects

For trichotillomania (F2,974.04=95.79, P<.001), RBFBS scores decreased from a mean of 7.84 (SD 2.29) to a mean of 6.19 (SD 2.68) at week 5-7 (-1.65 points, 18.32%; Hedges g=0.66, 95% CI [0.57, 0.75], n=543), and to a mean of 5.27 (SD 2.73) at week 14-16 (-2.56 points, 29.90%; Hedges g=1.01, 95% CI [0.88, 1.14], n=422). For excoriation disorder (F2,955.75=321.99, P<.001), RBFBS scores decreased from a mean of 7.89 (SD 2.14) to a mean of 6.34 (SD 2.51) at week 5-7 (-1.55 points, 17.51%; Hedges g=0.66, 95% CI [0.57, 0.75], n=528), and to a mean of 5.13 (SD 2.57) at week 14-16 (-2.74 points, 32.79%; Hedges g=1.16, 95% CI [1.02, 1.30], n=409).

### Age Group Differences

For trichotillomania, there was a significant main effect of time (F(2, 974.04)=95.79, p< .001), a significant effect of age category (F(2, 534.61)=17.43, p< .001), and a significant age by time interaction (F(4, 974.37)=2.66, p=.032). For excoriation disorder, there was a significant main effect of time (F(2, 955.75)=321.99, p< .001), age category (F(2, 531.26)=19.58, p< .001), and age by time interaction (F(4, 954.88)=3.45, p=.008). At baseline in the trichotillomania sample, adults showed higher scores than both adolescents (difference = 0.809, p=.028) and children (difference = 2.249, p<.001). Similarly at baseline in the excoriation sample, adults showed higher scores than both adolescents (difference =1.52, p< .001) and children (difference =1.94, p< .001). Treatment effects varied across age groups for both conditions (trichotillomania: adults g=1.11 [0.96, 1.27], adolescents g=0.81 [0.46, 1.15], children g=0.78 [0.47, 1.09]; excoriation: adults g=1.23 [1.07, 1.39], adolescents g=0.68 [0.33, 1.03], children g=1.54 [0.99, 2.09]; see Table 2 for means and sample sizes by timepoint). Overall, while all age groups showed meaningful improvement, both conditions showed age-related differences in baseline severity and treatment response, with adults generally showing higher baseline severity and strong treatment effects across both conditions.

**Table 2.** RBFBS Scores by Age Group, Condition, and Time Point.

| Age Group |  | Trichotillomania |  |  | Excoriation |  |  |
| --- | --- | --- | --- | --- | --- | --- | --- |
|  |  | Baseline | Week 5-7 | Week 14-16 | Baseline | Week 5-7 | Week 14-16 |
| Children | n(%) | 57 (100.00) | 57 (100.00) | 47 (82.46) | 40 (100.00) | 40 (100.00) | 30 (75.00) |
|  | M (SD) | 5.95 (2.49) | 4.93 (2.70) | 4.11 (2.55) | 6.22 (2.01) | 5.42 (2.33) | 3.33 (2.04) |
| Adolescents | n(%) | 75 (100.00) | 75 (100.00) | 62 (82.67) | 46 (100.00) | 46 (100.00) | 35 (76.09) |
|  | M (SD) | 7.39 (2.32) | 6.25 (2.61) | 5.21 (2.80) | 6.65 (2.24) | 5.46 (2.71) | 4.54 (2.82) |
| Adults | n(%) | 408 (100.00) | 408 (100.00) | 311 (76.23) | 442 (100.00) | 442 (100.00) | 344 (77.83) |
|  | M (SD) | 8.20 (2.11) | 6.36 (2.66) | 5.47 (2.70) | 8.17 (2.03) | 6.52 (2.47) | 5.35 (2.52) |

Note: M = Mean; RBFBS = Repetitive Body Focused Behavior Scale; SD = Standard Deviation
Age category data was missing for 3 trichotillomania members (0.6% of sample) who otherwise met all inclusion criteria. These participants had baseline scores M=6.67 (SD=2.08), midpoint scores M=6.00 (SD=0.00), and for the 2 who completed the endpoint assessment, M=4.00 (SD=4.24).

### Individual-Level Changes and Response Rates

For trichotillomania, median RBFBS hair score improvement was 33.33% (IQR: 11.11% to 54.55%) at week 14-16, with week 5-7 median improvement at 14.29% (IQR: 0.0% to 38.75%). Among those with week 14-16 data, 33.2% (140/422) had ≥45% reduction, 44.1% (186/422) had ≥35% reduction, 60.9% (257/422) had ≥25% reduction, 90.5% (382/422) had ≥0% reduction, and 9.5% (40/422) showed worsening of symptoms.

For excoriation disorder, median RBFBS score improvement was 33.33% (IQR: 12.50%-57.14%), with week 5-7 median improvement at 14.29% (IQR: 0.00%-37.50%). Among those with week 14-16 data, 37.2% (152/409) had ≥45% reduction, 48.7% (199/409) had ≥35% reduction, 63.3% (259/409) had ≥25% reduction, 90.5% (370/409) had ≥0% reduction, and 9.5% (39/409) showed worsening of symptoms.

### Depression, Anxiety, and Stress, Quality of Life and Disability

Both trichotillomania and excoriation disorder groups showed significant improvements in psychological symptoms, with small to moderate effects for depression (g=0.26-0.29), anxiety (g=0.22-0.26), and stress (g=0.27-0.28). Similar improvements were observed in functional outcomes, including quality of life (g=-0.26-0.25) and disability (g=0.25-0.30) (all Ps<.001; see Table 3 for means, Supplemental Figure 1-2 for changes over time, and Supplementary Results for detailed analyses).

**Figure 1.**
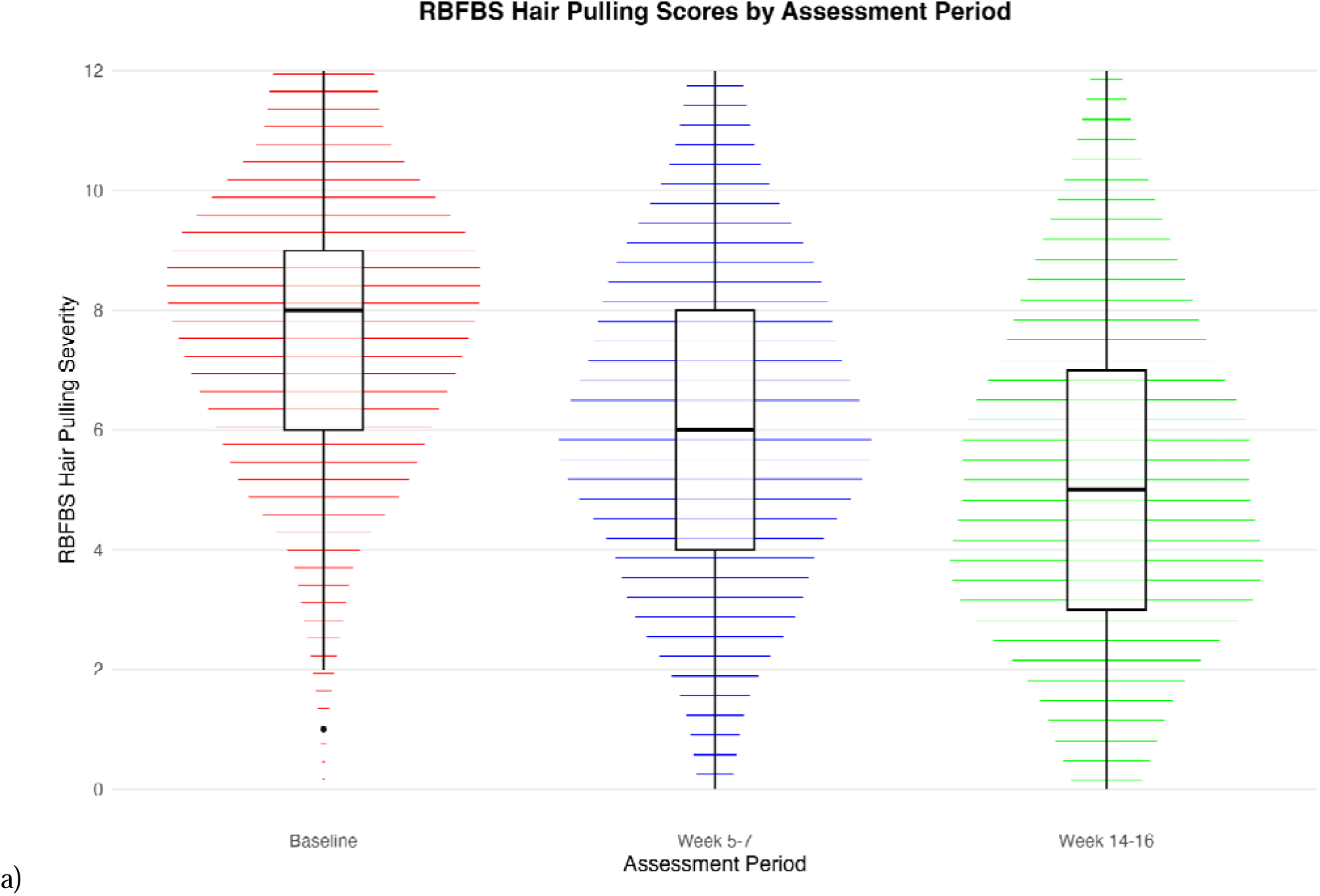
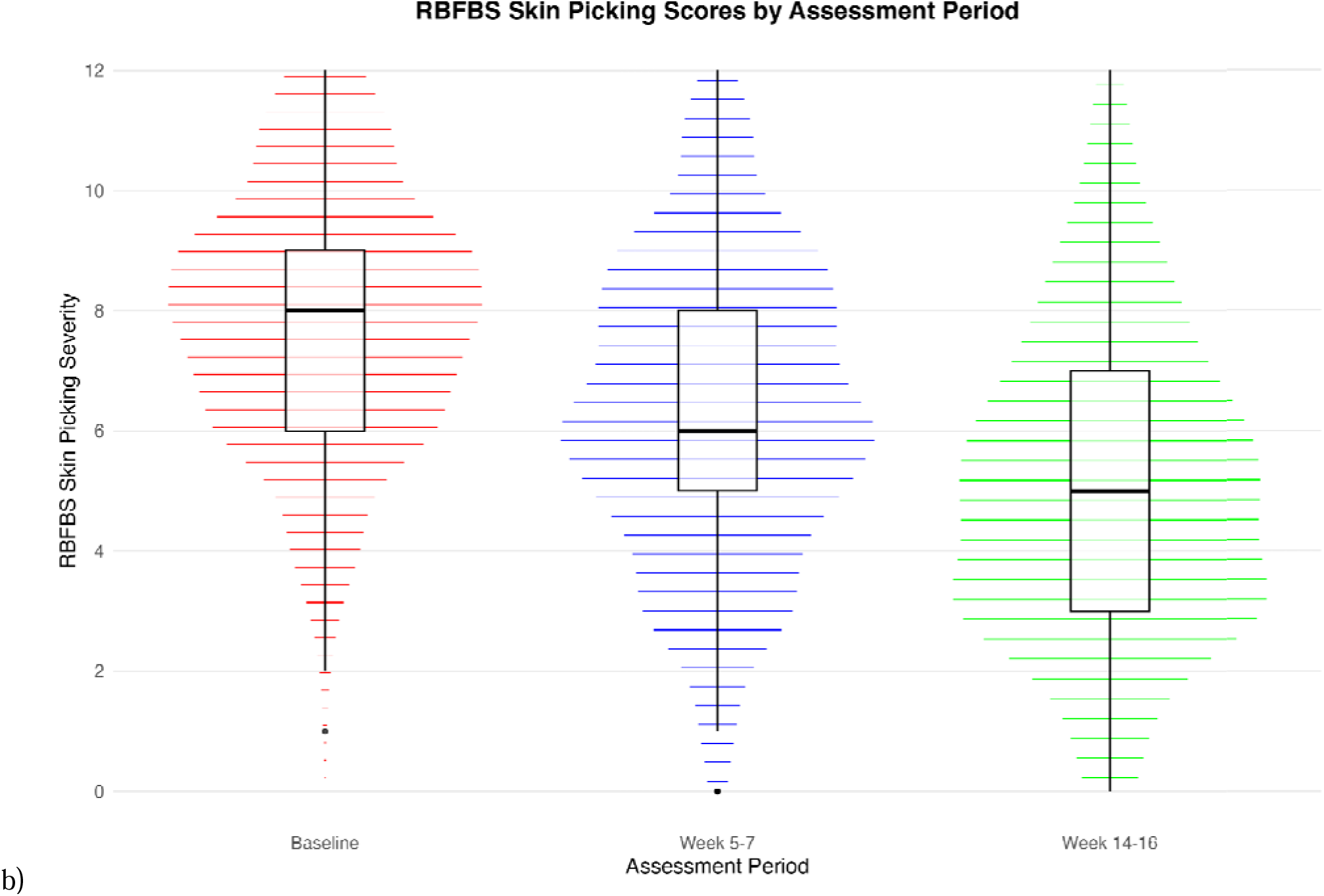
RBFBS Scores by Assessment Period. Note: RBFBS = Repetitive Body Focused Behavior Scale Changes in a) trichotillomania (top) and b) excoriation (bottom) symptoms as assessed by the RBFBS with treatment. Median and IQRs are indicated in the box-and-whisker plots. *P*<.001 for the effect of assessment period.

**Table 3.** Clinical assessments by treatment time point.

| Outcome scale and assessment time point | Trichotillomania |  |  | Excoriation |  |  |
| --- | --- | --- | --- | --- | --- | --- |
|  | Valid, n | Missing*, n | Mean (SD) | Valid, n | Missing*, n | Mean (SD) |
| **RBFB<sup>a</sup>** |  |  |  |  |  |  |
| Baseline | 543 | 0 | 7.84 (2.29) | 528 | 0 | 7.89 (2.14) |
| Week 5-7 | 543 | 0 | 6.19 (2.68) | 528 | 0 | 6.34 (2.51) |
| Week 14-16 | 422 | 121 | 5.27 (2.73) | 409 | 119 | 5.13 (2.57) |
| Week 17-28 | 365 | 178 | 4.83 (2.79) | 345 | 183 | 4.59 (2.31) |
| Week 29-40 | 229 | 314 | 4.51 (2.59) | 205 | 323 | 4.40 (2.07) |
| Week 41-52 | 122 | 421 | 4.23 (2.50) | 134 | 394 | 4.40 (2.32) |
| **DASS<sup>b</sup> depression** |  |  |  |  |  |  |
| Baseline | 488 | 55 | 9.47 (8.17) | 491 | 37 | 9.99 (8.84) |
| Week 5-7 | 508 | 35 | 8.42 (8.17) | 505 | 23 | 8.83 (8.08) |
| Week 14-16 | 406 | 137 | 7.26 (7.64) | 398 | 130 | 7.43 (7.28) |
| **DASS anxiety** |  |  |  |  |  |  |
| Baseline | 488 | 55 | 7.19 (7.05) | 491 | 37 | 7.54 (7.22) |
| Week 5-7 | 508 | 35 | 6.59 (6.97) | 505 | 23 | 6.74 (6.42) |
| Week 14-16 | 406 | 137 | 5.65 (6.30) | 398 | 130 | 5.86 (6.01) |
| **DASS stress** |  |  |  |  |  |  |
| Baseline | 488 | 55 | 14.41 (8.27) | 491 | 37 | 14.66 (8.09) |
| Week 5-7 | 508 | 35 | 13.35 (8.41) | 505 | 23 | 13.98 (7.70) |
| Week 14-16 | 406 | 137 | 11.96 (8.10) | 398 | 130 | 12.30 (7.53) |
| **QLESQ<sup>c</sup>** |  |  |  |  |  |  |
| Baseline | 390 | 153 | 63.19 (13.29) | 421 | 107 | 60.49 (14.98) |
| Week 5-7 | 402 | 141 | 65.10 (13.62) | 433 | 95 | 62.82 (15.08) |
| Week 14-16 | 309 | 234 | 67.07<br>(15.04) | 341 | 187 | 65.14 (14.51) |
| **WHODAS<sup>d</sup>** |  |  |  |  |  |  |
| Baseline | 382 | 161 | 7.98 (6.02) | 417 | 111 | 9.35 (7.32) |
| Week 5-7 | 398 | 145 | 7.15 (6.09) | 427 | 101 | 8.77 (7.38) |
| Week 14-16 | 305 | 238 | 6.03 (6.37) | 338 | 190 | 7.30 (6.69) |

\*Missing n's reflect patients who discontinued or completed treatment prior to that assessment timepoint. Treatment completion could occur due to meeting treatment goals, premature discontinuation, or switching to treating another condition as primary. Due to the naturalistic study design, we could not reliably differentiate between these reasons for their treatment ending. For weeks 17-52, missing n's represent those who did not participate in optional follow-up assessments after the active treatment phase (weeks 1-16).
<sup>a</sup>RBFB: Repetitive Body Focused Behavior Scale
<sup>b</sup>DASS: Depression Anxiety and Stress Scale—21
<sup>c</sup>QLESQ: Quality of Life Enjoyment and Satisfaction Questionnaire—Short Form
<sup>d</sup>WHODAS = World Health Organization Disability Assessment Schedule (2.0)

### Longitudinal follow-up

Treatment gains were maintained or improved during follow-up periods through week 52 (trichotillomania: F(5,1812.16)=212.27, P<.001; excoriation: (F(5,1425.68)=238.34, P<.001). For trichotillomania, mean symptom reduction from baseline was 38.8% at weeks 17-28 (n=365; Hedges g=1.18 [95% CI: 1.03-1.34]), 42.2% at weeks 29-40 (n=229; g=1.34 [95% CI: 1.14-1.55]), and 45.5% at weeks 41-52 (n=122; g=1.51 [95% CI: 1.23-1.79]). Similarly for excoriation, mean symptom reduction from baseline was 40.9% at weeks 17-28 (n=345; g=1.44 [95% CI: 1.27-1.61]), 42.9% at weeks 29-40 (n=205; g=1.59 [95% CI: 1.37-1.81]), and 44.1% at weeks 41-52 (n=134; g=1.56 [95% CI: 1.29-1.84]). See Figure 2 and Supplementary Results for detailed longitudinal analyses.

**Figure 2.**
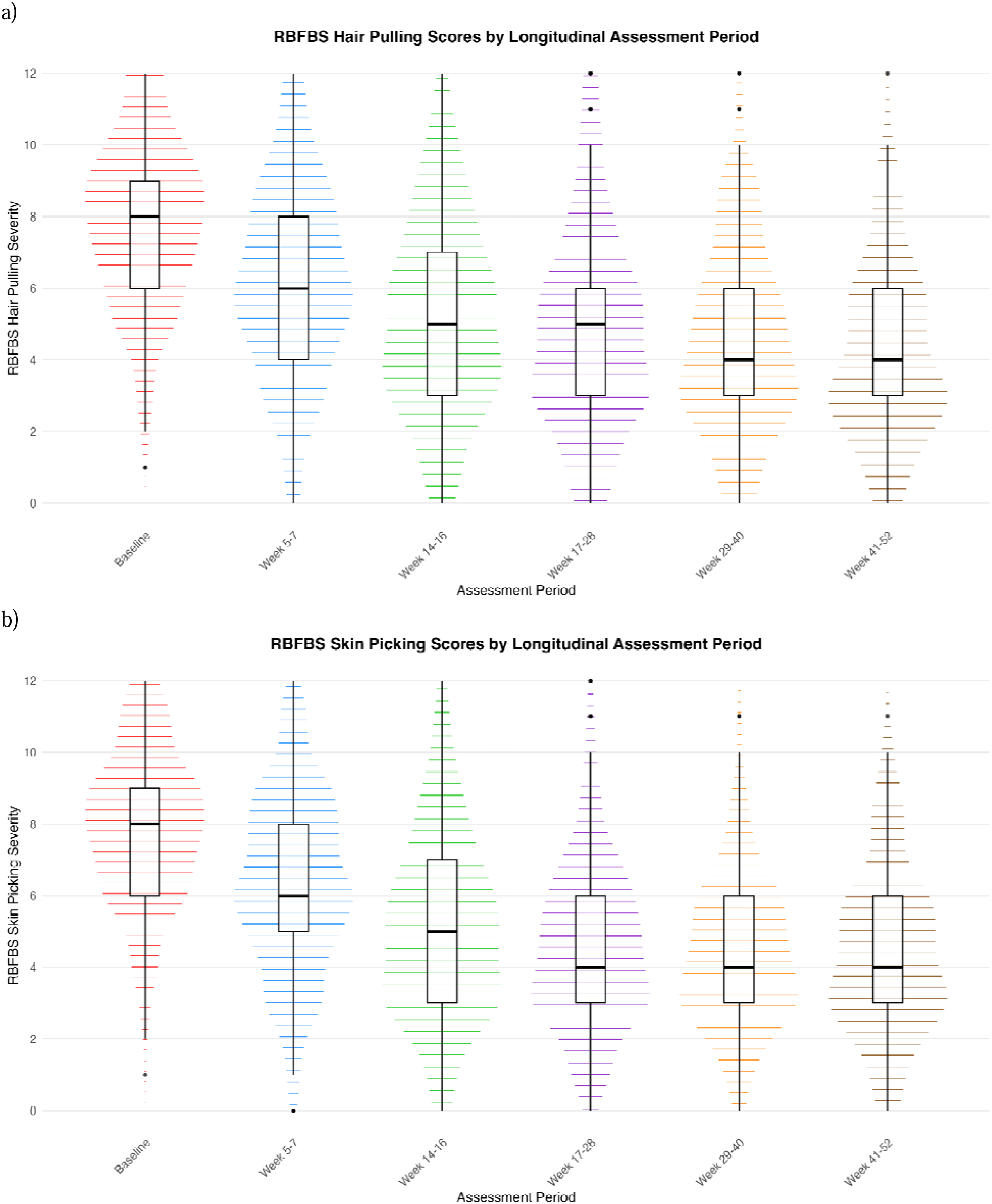
Longitudinal RBFBS Scores by Assessment Period. Note: RBFBS = Repetitive Body Focused Behavior Scale Changes in a) trichotillomania (top) and b) excoriation (bottom) symptoms as assessed by the RBFBS with treatment through 52-week follow-up. Median and IQRs are indicated in the box-and-whisker plots. P<.001 for the effect of assessment period.

## Discussion

This retrospective observational study demonstrates that HRT delivered via virtual therapy is effective for treating both trichotillomania and excoriation disorder in a large naturalistic sample. Treatment resulted in significant symptom reduction, with trichotillomania and excoriation decreasing by a mean of 29.9% and 32.8% respectively (g=1.01 and 1.16). Moreover, 44.1% of trichotillomania patients and 48.7% of excoriation disorder patients achieved ≥35% reduction in symptoms. Treatment also resulted in significant improvements in commonly co-occurring symptoms of depression, anxiety, and stress (g=0.22-0.29), and both groups demonstrated improvements in quality of life (g= -0.26-0.25) and disability (g=0.25-0.30). These secondary outcome improvements suggest that targeted BFRB treatment can lead to broader functional recovery. Importantly, these improvements were not only maintained but continued to strengthen during extended follow-up periods through week 52. While sample sizes decreased during follow-up, as expected in a naturalistic setting, both conditions showed sustained or enhanced treatment gains, suggesting potential long-term benefits of virtual therapy-delivered HRT. These findings from a large-scale, real-world setting provide promising evidence for virtual therapy-delivered HRT as a treatment option for these historically under-treated conditions. This is particularly important given that BFRBs typically onset around age 11^14^ and remain chronic if untreated, with only 50% of trichotillomania patients^16^ and 87.1% of excoriation disorder patients ever accessing care^62^. Even when psychiatric referrals for BFRBs are made, the majority of patients do not successfully engage in treatment ^63^, highlighting the critical need for more accessible treatment options.

While outcomes were analyzed at standardized timepoints (weeks 5-7 and 14-16), treatment duration was determined by individual clinical needs. The mean treatment duration was 14.64±2.50 weeks (7.71±2.61 sessions) for trichotillomania and 14.54±2.69 weeks (7.73±2.68 sessions) for excoriation, which aligns with typical HRT protocols, which have shown significant symptom reduction across treatment periods ranging from 8 weekly sessions to several months ^33,36,64,65^. While treatment duration was comparable to in-person HRT, the virtual therapy format eliminated geographical barriers. It also decreases travel burden and may have been associated with lower out-of-pocket costs for individual patients through high insurance utilization (>80% of patients). This enhanced accessibility to specialized BFRB treatment particularly benefits patients in areas with limited access to HRT specialists.

Despite HRT’s established efficacy^26,27,66^, significant barriers limit treatment access, spurring the development of technology-enabled delivery methods. Initial internet-delivered self-help programs for trichotillomania ^67^ and excoriation disorder ^68^ showed initial promise, but demonstrated modest outcomes in controlled trials ^69^. More recent studies of video-based teletherapy show promising outcomes. A randomized trial demonstrated significant reductions in hair-pulling severity in adults, with improvements maintained at 12-week follow-up ^43^. Similarly, a pilot study of video-delivered treatment for adolescents with trichotillomania showed significant symptom reduction with high treatment completion rates^44^. This evolution in delivery method is especially valuable as individuals with BFRBs report openness to internet-delivered interventions,^70^ potentially addressing barriers related to shame and access.^71^

Our findings align with emerging evidence for technology-enabled BFRB treatment. A recent large-scale evaluation of internet-based therapist-assisted programs (N=1,423) found significant symptom reduction for both trichotillomania and excoriation disorder (d=0.82 and d=0.99, respectively)^72^. While their treatment model shared some features with our approach—including licensed therapist support and integration of HRT with other evidence-based components—our video-based structured sessions used a different delivery format than their self-paced, chat-based approach. Our platform incorporated real-time video sessions allowing therapists and patients to interact while practicing exposures and exercises in typical environments where symptoms often occur. In-app messaging demonstrated high engagement (99-100% of patients sent messages, 97-99% sent 10+ messages), while symptom tracking tools and a moderated online community provided supplementary support that may have helped reduce social isolation commonly experienced with BFRBs. This approach thus combined effective HRT delivery with robust between-session engagement tools to address clinical and support needs.

Treatment outcomes were consistent across age groups. While adults initially showed higher severity scores in both conditions, endpoint differences remained only between adults/adolescents and children. These findings demonstrate that virtual therapy HRT can be effectively adapted across developmental stages. This is particularly important since BFRB symptoms typically onset in childhood/early adolescence^14^, yet treatment is often delayed for years. Early virtual therapy intervention across age groups could help prevent the progression to more severe or chronic symptoms that occur with delayed treatment.

Several limitations warrant discussion. The observational design (thus, without a control group or randomization) precludes causal inferences about treatment effects, although our findings align with previous controlled HRT trials. A constraint was using the RBFBS as our primary outcome measure instead of the Generic BFRB Scale-8 (GBS-8) ^54^ or the Massachusetts General Hospital Hairpulling Symptom Severity Scale (MGH-HS) ^53^. While the RBFBS parent-report shows strong psychometric properties in youth samples and the adult version demonstrates promising convergent validity with the GBS-8, the adult self-report version lacks comprehensive psychometric validation (including discriminant validity and internal consistency), and limits our ability to make direct comparisons with previous studies. Additional limitations include a lack of formal treatment fidelity assessment (despite standardized training and supervision) and missing data, which reflects typical real-world treatment engagement.

In conclusion, virtual therapy-delivered HRT demonstrated clinically significant improvements in both trichotillomania and excoriation disorder symptoms across severity levels and age groups, while also improving depression, anxiety, stress, and quality of life outcomes in this large naturalistic sample. High engagement and strong insurance utilization suggest this treatment approach offers a scalable, accessible option for evidence-based care that could reduce the substantial gap between symptom onset and treatment typically seen in these conditions.

## Data Availability

The data sets analyzed during this study are proprietary business assets of NOCD Inc., and are not publicly available. The data may be available from the corresponding author on reasonable request with appropriate data use agreements in place.

## Abbreviations

ACT: Acceptance and Commitment Therapy
ADHD: Attention Deficit Hyperactivity Disorder
AEBT: Acceptance-Enhanced Behavior Therapy
BFRB: Body-Focused Repetitive Behavior
CI: Confidence Interval
DASS-21: Depression, Anxiety, and Stress Scales
DBT: Dialectical Behavior Therapy
DIAMOND: Diagnostic Interview for Anxiety, Mood, and Obsessive-Compulsive and Related Neuropsychiatric Disorders
ED: Excoriation Disorder
GBS-8: Generic Body-Focused Repetitive Behavior Scale-8
HRT: Habit Reversal Training
IQR: Interquartile Range
MGH-HS: Massachusetts General Hospital Hairpulling Symptom Severity Scale
OCD: Obsessive-Compulsive Disorder
QLESQ-SF: Quality of Life Enjoyment and Satisfaction Questionnaire-Short Form
RBFBS: Repetitive Body Focused Behavior Scale
RBFBS-p: Repetitive Body Focused Behavior Scale-Parent Report
RBFBS-SR: Repetitive Body Focused Behavior Scale-Self Report
SD: Standard Deviation
SNRI: Serotonin-Norepinephrine Reuptake Inhibitor
SSRI: Selective Serotonin Reuptake Inhibitor
TCA: Tricyclic Antidepressant
WHODAS 2.0: World Health Organization Disability Assessment Schedule 2.0

## Relevant Financial Relationships

JDF, CCB, PBM, NRF, MN, LT, SMS, AR, NL, and RM report personal fees from NOCD Inc. CM has no relevant financial relationships to disclose.

## Acknowledgments

The authors thank the NOCD therapists and Member Advocates for their help in facilitating treatment and care experiences.

