## SupplementalFiles for "Virtual Therapy Habit Reversal Training for Body-Focused Repetitive Behaviors: Clinical Outcomes from a Large Real-World Sample of Youth and Adults" (from another version of this preprint: version 1, 2025-03-14)

**Supplementary Materials**

**Methods**

*Initial Evaluation and Clinical Assessments*

Patients entered treatment through NOCD's standard clinical pathway, either through self-referral or referral from insurance providers or medical professionals. Initial diagnostic evaluations were conducted by NOCD-trained therapists over two sessions, comprising a comprehensive biopsychosocial assessment and a semi-structured diagnostic interview using the Diagnostic Interview for Anxiety, Mood, and Obsessive-Compulsive and Related Neuropsychiatric Disorders (DIAMOND; ^1,2^) Individuals were eligible for treatment if they met DSM-5 criteria for trichotillomania or excoriation disorder as their primary diagnosis. NOCD typically serves individuals aged 5 and older.

Individuals receiving an "extreme" rating on the DIAMOND clinician-rated severity scale were generally referred to more intensive treatment options (e.g., intensive outpatient programs, partial hospitalization programs, or residential treatment). Exceptions (n=2 with excoriation disorder in this cohort) were considered based on clinical judgment, taking into account factors such as treatment motivation, presence of comorbidities, and availability of family support. Additional referrals were made as needed for concurrent psychiatric or substance use concerns that might interfere with HRT treatment.

*Therapist Qualifications and Training*

Therapists completed an intensive 12 week training on the treatment of OCD and obsessive-compulsive related disorders, as well as commonly co-occurring conditions. Training specific to treatment of BFRBs was delivered in a hybrid model consisting of both live and asynchronous didactic learning with role play demonstrations followed by a post-test to ascertain knowledge and skills accrued. Therapists attended consultation groups to receive ongoing guidance on treatment delivery, as well as to ask case-specific questions.

*Assessment Procedures*

Patients completed standardized assessments at three primary timepoints: baseline, weeks 5-7, and weeks 14-16. These assessment windows were chosen based on typical HRT response trajectories, with weeks 5-7 capturing early treatment response and weeks 14-16 representing a clinically meaningful endpoint. When multiple assessments were completed within a timepoint window (e.g., both week 5 and week 6), we selected the most recent assessment for analysis. Beyond week 16, additional assessments were collected during follow-up maintenance periods (weeks 17-28, 29-40, and 41-52) to evaluate longer-term outcomes. Treatment duration and frequency during this maintenance phase varied based on individual clinical needs, with some patients requiring more intensive ongoing support while others transitioned to less frequent maintenance sessions.

*Secondary Outcome Measures*

1. Depression, Anxiety, and Stress Scale (DASS-21)^3^: 21-item self-report measure with three 7-item subscales assessing depression, anxiety, and stress. Items are rated from 0 ("Did not apply to me at all") to 3 ("Applied to me very much"), with subscale scores multiplied by 2 to yield final scores ranging from 0-42, where higher scores indicate greater symptom severity.
2. Quality of Life Enjoyment and Satisfaction Questionnaire-Short Form (Q-LES-Q-SF)^4^: 16 item self-report measure with the first 14 items scored on a 5-point scale and summed to yield a total percentage score ranging from 0-100, where higher scores indicate better quality of life.
3. World Health Organization Disability Assessment Schedule 2.0 (WHODAS 2.0)^5^: 12 item self-report measure assessing disability and functioning related to medical and/or mental health conditions. Items are scored from 0 (no difficulty) to 4 (extreme difficulty), with total scores ranging from 0-48. Higher scores indicate greater impairment.

*Data Analysis*

*Missing Data Analysis*. Missing data patterns were analyzed to evaluate Missing Completely at Random assumptions by comparing patients with weeks 14-16 data to those with only weeks 5-7 data on demographic and clinical characteristics using t-tests for continuous variables (age, baseline BFRD severity, baseline depression, anxiety, and stress scores) and chi-square tests for categorical variables (age category, gender). Missing data were handled by specifying whether missing cases were due to BFRB treatment ending versus non-completion of assessment forms while still in treatment.

*Satisfaction Analysis.* Treatment satisfaction was analyzed through descriptive statistics and by testing its relationship with outcomes using correlation analysis and mixed effects models.

**Results**

*Missing Data Analysis*

For trichotillomania, comparisons between those who provided versus did not provide week 14-16 assessments showed no significant differences in age category (χ²(1)=1.97, P=.160), gender (χ²(2)=4.65, P=.098), or baseline BFRD severity (t(190.03)=-0.11, P=.916), with effect sizes being negligible (Cohen's d=-0.01). Similarly for excoriation disorder, there were no significant differences in age category (χ²(1)=0.10, P=.753), gender (χ²(2)=0.78, P=.676), or baseline BFRD severity (t(173.89)=-0.45, P=.650) between those who provided versus did not provide week 14-16 assessments, with all effect sizes being negligible (|d|<.2). These findings support Missing Completely at Random assumptions for both conditions.

**Supplementary Table S1. RBFBS Assessment Completion and Treatment Duration by Disorder and Time Point**

|  | **Trich** | | | | **Excoriation** | | | |
| --- | --- | --- | --- | --- | --- | --- | --- | --- |
|  | ***Weeks 5-7 only*** | | ***Week 14-16*** | | ***Weeks 5-7 only*** | | ***Week 14-16*** | |
| n (%)^1^ | 121 (22.3) | | 422 (77.7) | | 119 (22.5) | | 409 (77.5) | |
|  | M (SD) | Med (IQR) | M (SD) | Med (IQR) | M (SD) | Med (IQR) | M (SD) | Med (IQR) |
| Age | 27.42 (11.04) | 27 (20-34) | 26.29 (11.09) | 25 (17-33) | 29.08 (12.36) | 28 (21-36) | 27.54 (10.80) | 26 (21-32) |
| Treatment Duration^1^ (weeks) | 10.39 (1.98) | 11 (9-12) | 31.54 (12.54) | 30 (20-43) | 10.01 (2.22) | 10 (9-12) | 31.49 (13.15) | 29 (19-45) |
| Number of visits | 6.65 (1.58) | 6 (5-7) | 14.43 (7.02) | 13 (9-19) | 6.34 (1.38) | 6 (5-7) | 14.90 (7.78) | 13 (9-19) |

^1^Note: For trichotillomania, the sample includes 543 patients who completed baseline and at least one follow-up assessment. The "Weeks 5-7 only" group consists of patients who discontinued treatment before the week 14-16 assessment. Those in the "Week 14-16" group remained engaged throughout the primary assessment period.

^2^Treatment duration represents the full course of treatment, including maintenance sessions after week 16 for those who continued. Week 14-16 completers had longer treatment durations because they remained engaged in treatment for the entire period.

Note: M = Mean; SD = Standard Deviation; Med = Median; IQR = Interquartile Range

*Treatment Engagement and Satisfaction*

*App Use and Messaging.* For trichotillomania, 542/543 (99.82%) patients sent at least one in-app message to their therapist and 535/543 (98.53%) sent 10 or more messages, with a mean of 81.40 (SD 58.28) app opens and 68.62 (SD 56.00) messages. For excoriation disorder, 526/528 (99.62%) patients sent at least one message and 514/528 (97.35%) sent 10 or more messages, with a mean of 83.93 (SD 80.67) app opens and 71.31 (SD 67.69) messages.

*Treatment Satisfaction.* Treatment satisfaction ratings were high for both conditions on a 0-5 Likert scale. For trichotillomania, among the 335 patients who provided satisfaction ratings (61.7% of sample), mean satisfaction was 4.77 (SD=0.51; median=5.0, IQR=4.87-5.00), and adding satisfaction to the mixed effects model did not significantly improve fit (χ²(3)=7.16, P=.07), with no significant correlation between satisfaction and symptom improvement (r=-0.041, P=.50). Similarly for excoriation disorder, among the 333 patients who provided satisfaction ratings (63.1% of sample), mean satisfaction was 4.76 (SD=0.58; median=5.0, IQR=4.94-5.00), and adding satisfaction did not improve model fit (χ²(3)=9.71, P=.02), with no significant correlation between satisfaction and symptom improvement (r=-0.11, P=.07). For both conditions, the lack of relationship between satisfaction and outcomes should be interpreted in the context of consistently high satisfaction ratings with limited variability, suggesting a potential ceiling effect.

*Age Group Differences in Primary Outcome*

Linear mixed models revealed similar patterns between conditions. For trichotillomania, there was a significant main effect of time (F(2,985.9)=245.58, P<.001) and a significant effect of age category (F(2,534.61)=17.43, P<.001), with a significant age by time interaction (F(4,974.37)=2.66, P=.032). For excoriation disorder, there were significant main effects of time (F(2,955.75)=321.99, P<.001) and age category (F(2,531.26)=19.58, P<.001), and a significant age by time interaction (F(4,954.88)=3.45, P=.008).

At baseline in the trichotillomania sample, adults (M=8.20, SD=2.11) showed significantly higher scores than adolescents (M=7.39, SD=2.32; difference=0.81, P=.028) and children (M=5.95, SD=2.49; difference=2.25, P<.001). Adolescents also showed significantly higher scores than children (difference=1.44, P=.003). Similarly, at baseline in the excoriation sample, adults showed significantly higher scores than both adolescents (difference=1.52, P<.001) and children (difference=1.94, P<.001).

Effect size calculations revealed improvements across all age groups. For trichotillomania, effects were large for adults (Hedges g=1.11, 95% CI [0.96, 1.27], n=311) and moderate for adolescents (Hedges g=0.81, 95% CI [0.46, 1.15], n=62) and children (Hedges g=0.78, 95% CI [0.47, 1.09], n=47). For excoriation disorder, effects were large for adults (Hedges g=1.23, 95% CI [1.07, 1.39], n=344) and children (Hedges g=1.54, 95% CI [0.99, 2.09], n=30), and moderate for adolescents (Hedges g=0.68, 95% CI [0.33, 1.03], n=35).

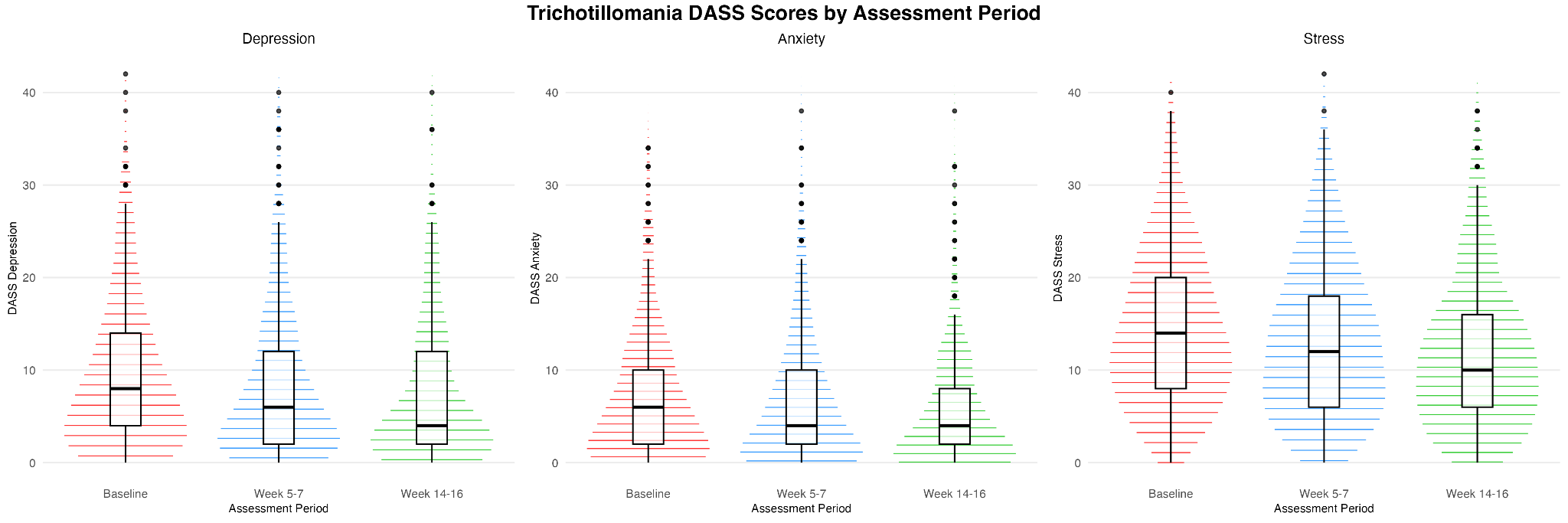

a)
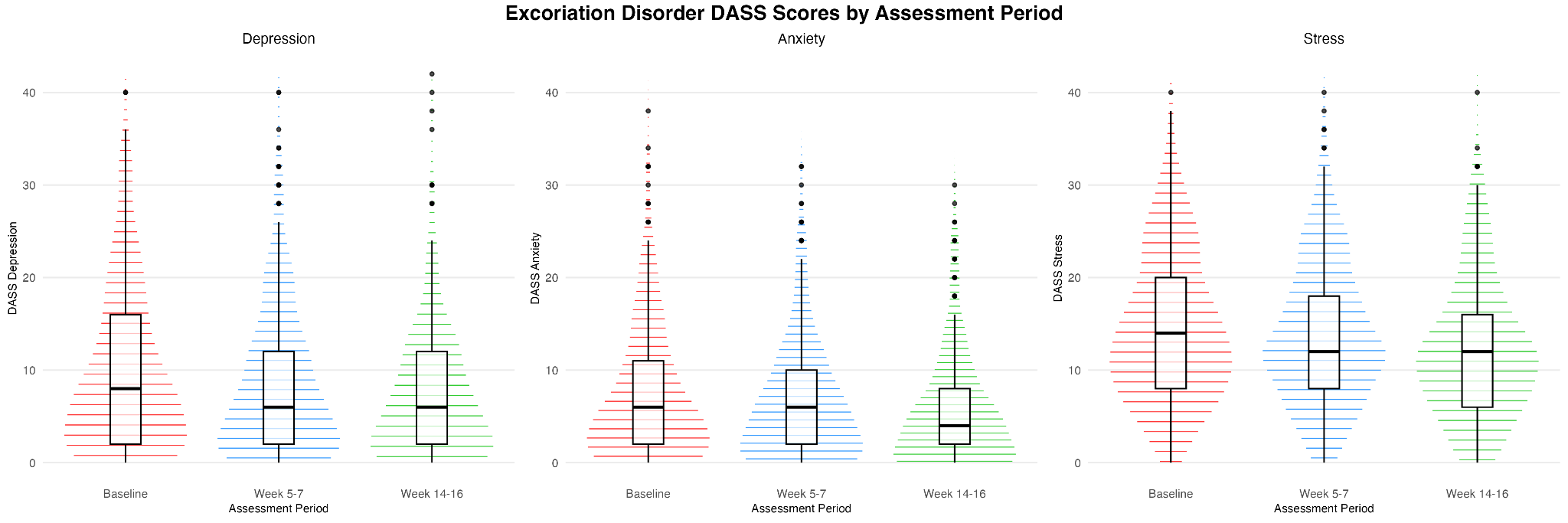

**Supplemental Figure S1.** Changes in depression, anxiety, and stress symptoms in a) trichotillomania (top) and b) excoriation (bottom) samples as assessed by the DASS with treatment. Median and IQRs are indicated in the box-and-whisker plots. *P*<.001 for the effect of assessment period.

a)
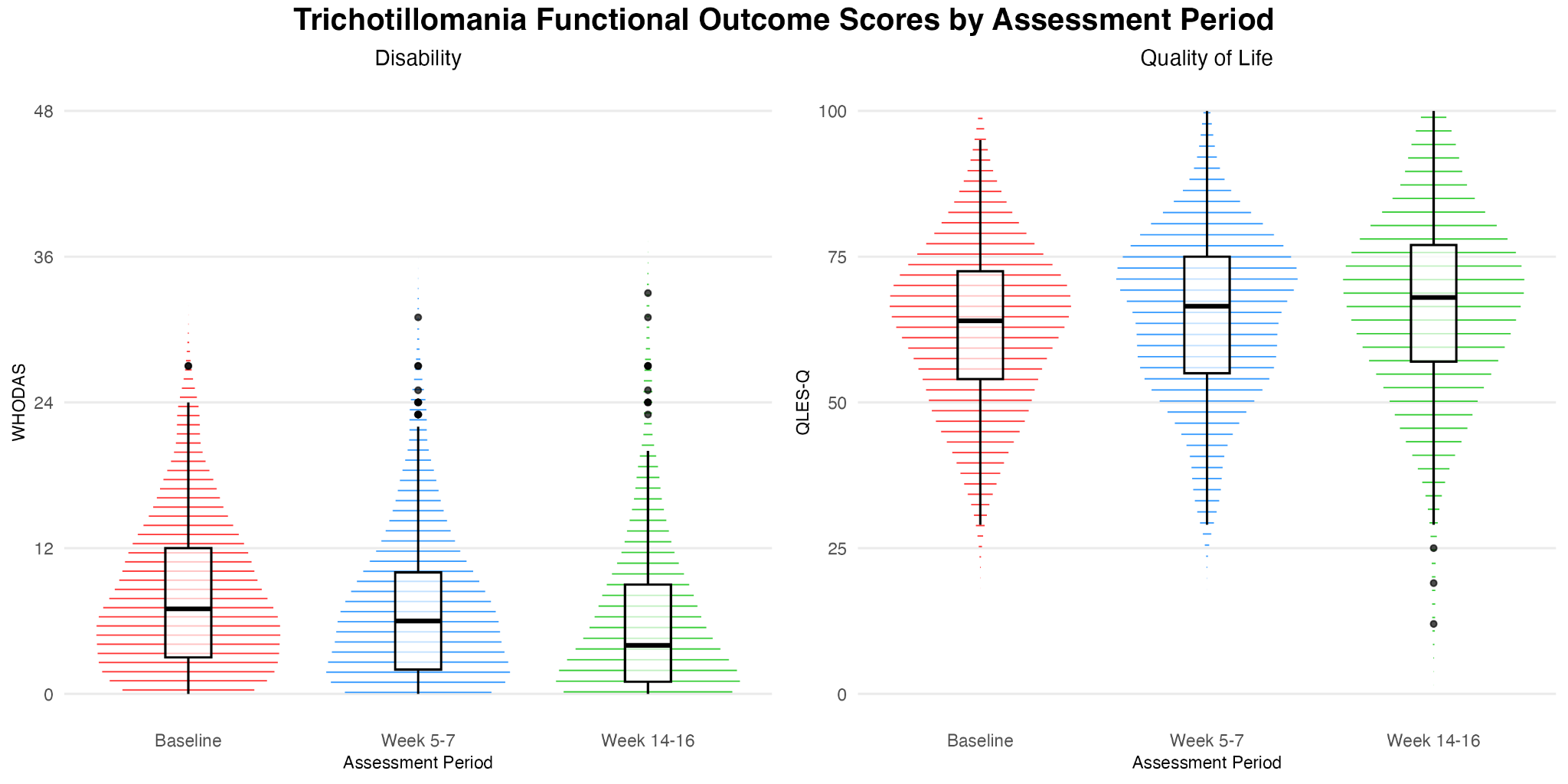

b)
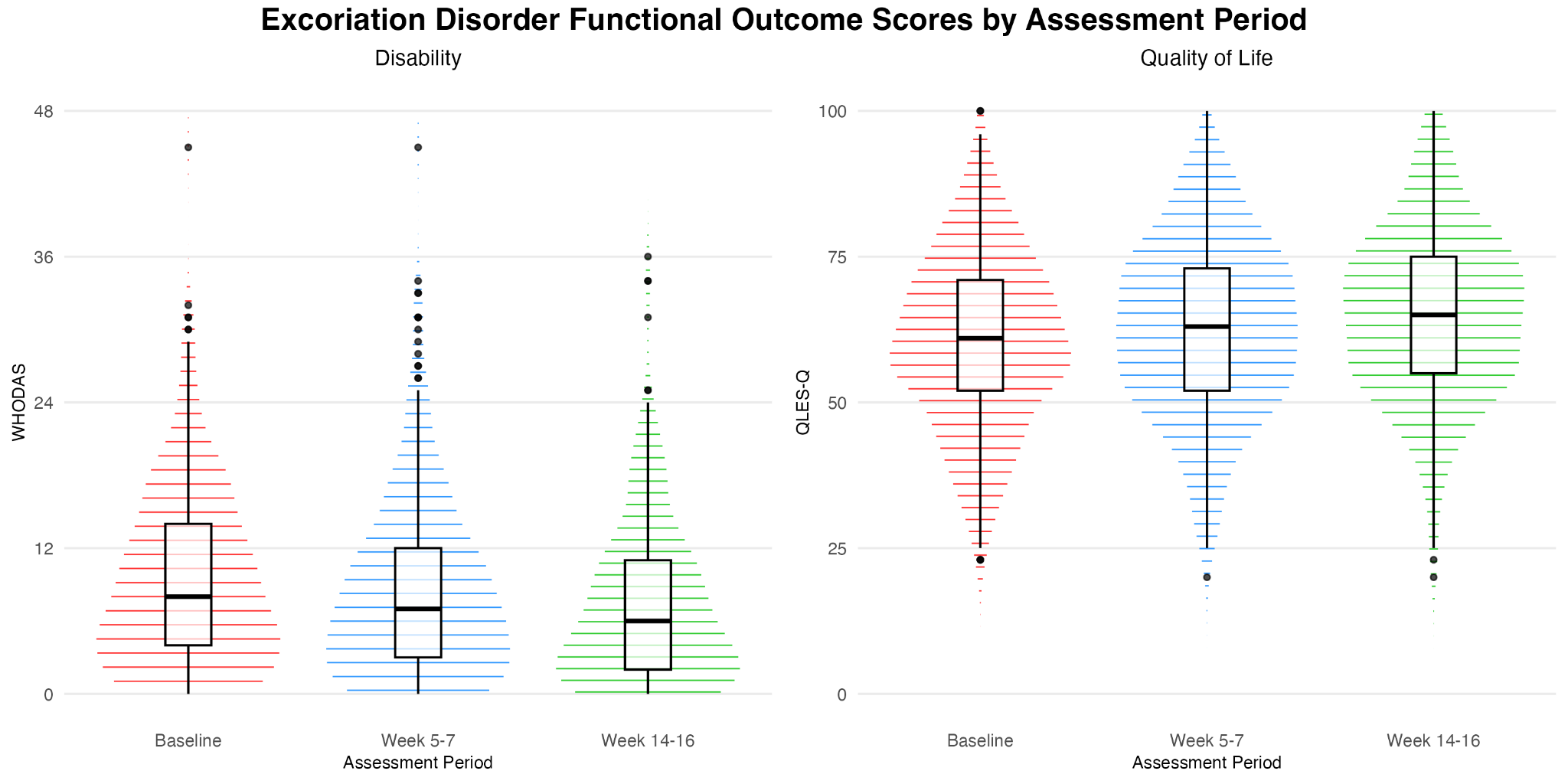

**Supplemental Figure S2.** Changes in disability and quality of life in a) trichotillomania (top) and b) excoriation (bottom) samples as assessed by the WHODAS and QLESQ with treatment. Median and IQRs are indicated in the box-and-whisker plots. *P*<.001 for the effect of assessment period.

*Depression, Anxiety, and Stress*

For trichotillomania, treatment resulted in significant improvements on the DASS depression (F(2,903.16)=24.77, P<.001; baseline to week 14-16 Hedges g=0.26, 95% CI [0.17, 0.36], n=384), DASS anxiety (F(2,906.9)=17.82, P<.001; baseline to week 14-16 Hedges g=0.22, 95% CI [0.13, 0.31], n=384), and DASS stress (F(2,903.31)=26.86, P<.001; baseline to week 14-16 Hedges g=0.28, 95% CI [0.18, 0.37], n=384) (see Supplemental Figure 1 and Table 3).

For excoriation disorder, significant improvements were also observed across DASS depression (F2,899.54=27.27, P<.001; baseline to week 14-16 Hedges g=0.29, 95% CI [0.19, 0.38]), DASS anxiety (F2,897.26=27.60, P<.001; baseline to week 14-16 Hedges g=0.26, 95% CI [0.18, 0.35]), and DASS stress (F2,900.34=24.23, P<.001; baseline to week 14-16 Hedges g=0.27, 95% CI [0.17, 0.36]) (see Supplemental Figure 1 and Table 3).

*Quality of Life and Disability*

For trichotillomania, treatment led to significant improvements in quality of life (F(2,709.85)=20.36, P<.001; baseline to week 14-16 Hedges g=-0.26, 95% CI [-0.36, -0.17], n=299) and disability (F(2,690.99)=30.35, P<.001; baseline to week 14-16 Hedges g=0.30, 95% CI [0.20, 0.40], n=293) (see Supplemental Figure 2 and Table 3).

For excoriation disorder, significant improvements were also observed in quality of life (F2,764.61=26.99, P<.001; baseline to week 14-16 Hedges g=-0.25, 95% CI [-0.34, -0.17]) and disability (F2,752.33=33.52, P<.001; baseline to week 14-16 Hedges g=0.25, 95% CI [0.17, 0.34]) (see Supplemental Figure 2 and Table 3).

*Longitudinal follow-up*

We conducted an analysis of longitudinal follow-up of BFRB severity scores after the active treatment period (weeks 1-16). For trichotillomania, at the follow-up windows of weeks 17-28, 29-40, and 41-52, most patients maintained their gains or made further improvements (effect of time: F(5,1812.16)=212.27, P<.001). From baseline to weeks 17-28 (n=365), hair pulling scores decreased from a mean of 7.89 to a mean of 4.83, representing a mean -3.06 (CI -3.36 to -2.76) point decrease (38.8%), Hedges g = -1.18, CI = -1.34 to -1.03. By weeks 29-40 (n=229), hair pulling scores improved to a mean of 4.51, representing a mean -3.29 (CI -3.65 to -2.93) point decrease (42.2%), Hedges g = -1.34, CI = -1.55 to -1.14. By weeks 41-52 (n=122), hair pulling scores improved to a mean of 4.23, representing a mean -3.52 (CI -4.04 to -3.01) point decrease (45.5%) from baseline, Hedges g = -1.51, CI = -1.79 to -1.23.

For excoriation, at the follow-up windows of weeks 17-28, 29-40, and 41-52, most patients maintained their gains or made further improvements (effect of time: F(5,1425.68)=238.34, P<.001). From baseline to weeks 17-28 (n=345), BFRB scores decreased from a mean of 7.77 to a mean of 4.59, representing a mean -3.17 (CI -3.44 to -2.90) point decrease (40.9%), Hedges g = -1.44, CI = -1.61 to -1.27. By weeks 29-40 (n=205), BFRB scores improved to a mean of 4.40, representing a mean -3.31 (CI -3.63 to -2.99) point decrease (42.9%), Hedges g = -1.59, CI = -1.81 to -1.37. By weeks 41-52 (n=134), BFRB scores improved to a mean of 4.40, representing a mean -3.48 (CI -3.91 to -3.05) point decrease (44.1%) from baseline, Hedges g = -1.56, CI = -1.84 to -1.29.
